# Cost-effectiveness and benefit-cost analyses of promoting handwashing with soap: a systematic review

**DOI:** 10.1101/2025.10.13.25337661

**Authors:** Ian Ross, David Bath, Joseph Wells, Robert Dreibelbis, Regina Ejemot-Nwadiaro, Joanna Esteves Mills, Giulia Greco, Catherine Pitt, Oliver Cumming

**Affiliations:** London School of Hygiene and Tropical Medicine, UK; Kampala International University, Uganda; University of Calabar, Nigeria; World Health Organization, Switzerland

## Abstract

**Background:** Promoting handwashing with soap reduces risk of diarrhoea by 30% and respiratory infections by 17%. Handwashing promotion in non-healthcare settings is widely considered cost-effective, but there is no systematic review on this topic. To inform resource allocation decisions, we reviewed the state and quality of evidence regarding cost-effectiveness and benefit-cost of interventions promoting handwashing with soap in domestic, educational, and childcare settings globally.

**Methods:** We searched Medline, Embase, Global Health, EconLit, and Web of Science for studies published to September 3, 2025, as well as grey literature (PROSPERO CRD42021288727). We included full economic evaluations comparing the cost of two or more interventions with their outcomes. We included interventions promoting the practice of handwashing with soap, including those providing information, motivational campaigns, and/or handwashing facilities. We scored quality of reporting using the Consolidated Health Economic Evaluation Reporting Standards.

**Results:** We identified 15 studies of which 3 were in high-income countries. Five used empirical data collection to evaluate interventions actually implemented and 10 modelled from secondary data only. Amongst the 3 medium- or high-quality studies reporting cost per disability-adjusted life-year averted, estimates ranged from US$ 37 to 937 (2024 prices). Of these 3 estimates, 2 were cost-effective compared to plausible thresholds for the respective country. In the only medium- or high-quality benefit-cost study, the mean benefit-cost ratio was 2.1 with “medium” levels of handwashing adoption (40% of population) and adherence (50% of those adopting). Few studies measured or modelled adoption of handwashing over time, and none which focused on diarrhoea also valued respiratory infections.

**Conclusion:** Promoting handwashing with soap is very likely to be cost-effective for interventions that successfully increase and sustain adoption of handwashing behaviours. More empirical studies are needed, especially those comparing multiple promotion options and valuing reductions in respiratory infections as well as diarrhoea.

## 1. Introduction

Poor hygiene contributes to a substantial disease burden, with 740,000 deaths from diarrhoea and respiratory infections in 2019 attributable to inadequate handwashing with soap (Wolf et al., 2023). Globally, only 26% of potential faecal contact events in domestic settings are followed by handwashing with soap, rising to 51% in regions with high access to handwashing facilities (Wolf et al., 2019). Improving levels of handwashing with soap could avoid morbidity and mortality, since its promotion is effective at reducing risk of diarrhoea by 30% (Wolf et al., 2022) and acute respiratory infections by 17% (Ross et al., 2023). It could also improve quality of life (e.g. feelings of pride or cleanliness) and reduce the direct costs of illness and associated productivity losses. However, the efficiency of handwashing investments remains unclear, particularly in a time of global health funding cuts.

Promoting handwashing with soap has long been considered cost-effective (Feachem, 1984). In healthcare settings, there is good evidence that interventions to improve hand hygiene are cost-effective or even cost-saving (Rice et al., 2023). For domestic settings, various iterations of the Disease Control Priorities study have ranked handwashing promotion amongst the most cost-effective interventions in any area of health (Horton, 2018; Laxminarayan et al., 2006). However, the economic evaluation evidence for domestic settings is thinner than for healthcare. A previous review across all of water, sanitation and hygiene (WASH)(Hutton & Chase, 2016) identified seven cost-effectiveness and benefit-cost analyses of promoting handwashing with soap; however, the review was not systematic, did not assess study quality, extracted few study characteristics, did not draw handwashing-specific conclusions, and is now a decade old.

To address these knowledge gaps, we assessed the state and quality of evidence regarding the cost-effectiveness and benefit-cost of interventions promoting handwashing with soap in domestic, school or childcare settings globally. Our findings will inform discussions around the forthcoming WHO guidelines on hand hygiene beyond healthcare settings, as well as the Lancet Commission on WASH and health (Amebelu et al., 2021). They can also inform improvements in the reporting quality of handwashing economic evaluations and the development of methodological guidelines.

## 2. Methods

### Study design

Our systematic review is reported according to PRISMA 2020 guidelines (Page et al., 2021), and was pre-registered with the International Prospective Register of Systematic Reviews (PROSPERO) - CRD42021288727. We followed best practice for systematic reviews of economic evaluation evidence (Mandrik et al., 2021).

### Search strategy

We searched Medline, Embase, Global Health, EconLit, Web of Science, Cochrane library, International Bibliography of the Social Sciences, Global Health Cost Effectiveness Analysis Registry, and the NHS Economic Evaluation Database, for literature published from January 1980 to September 3, 2025. We also searched for unpublished or grey literature studies using databases from National Bureau of Economic Research, International Initiative for Impact Evaluation, Research Papers in Economics, WHO Index Medicus (all regions), and Copenhagen Consensus Centre.

Our search strategy (Supplementary Material A) combines terms for economic evaluation and terms for promotion of handwashing with soap used in recent reviews (Ross et al., 2023; Wolf et al., 2022). We used Endnote (Clarivate, Philadelphia, USA) for de-duplication, Rayyan for managing blinded abstract screening (Ouzzani et al., 2016), and Microsoft Excel for data extraction. We also screened reference lists of included studies and previous reviews. Two reviewers (IR with DB or JW) independently screened titles, abstracts, and full texts. Differences between reviewers were reconciled by discussion, with recourse to a third reviewer (OC) if necessary.

### Selection criteria

Eligible settings were domestic (households), educational, or childcare, in any population worldwide. We excluded other settings, notably healthcare facilities, because they have fundamentally different disease transmission risk and target audience of interventions (usually trained staff rather than the public).

Eligible interventions promoted handwashing with soap through communication and/or provision of facilities or products. Examples of communication include mass media campaigns, door-to-door visits or group activities. Examples of provision include distribution or marketing of handwashing stations (fixed or mobile) or soap. Accordingly, we excluded studies without promotion, for example studies which assumed that populations spontaneously changed behaviour without external intervention. Following Ross et al. (2023), we included interventions combining handwashing with other interventions (e.g. water treatment, face masks) if handwashing was a “major” behavioural target of the intervention (Supplementary Material B). Following norms in the literature for disease burden (Wolf et al., 2023) and intervention effectiveness (Ejemot-Nwadiaro et al., 2021), we only included interventions promoting handwashing with soap and excluded those promoting alcohol-based hand rub, antimicrobial towels, or other soap alternatives.

Eligible study designs were full economic evaluations, i.e. comparisons of two or more options (one of which can be existing practice or doing nothing) in terms of their relative costs and outcomes (Drummond et al., 2015). Specifically we included benefit-cost analyses and cost-effectiveness analyses (included cost-utility analyses under this category). We excluded partial economic evaluations, such as those reporting cost analysis only (e.g. cost per person targeted/reached). We also excluded prospective economic appraisals, e.g. project planning documentation.

Eligible outcomes were summary measures of efficiency, such as a ratio (e.g. incremental cost-effectiveness ratio or benefit-cost ratio), a probability (e.g. that an intervention is cost-effective given a threshold), a difference (e.g. incremental net benefit), or net present value (Pitt et al., 2016). We also included studies that did not report such a measure but demonstrated dominance, e.g. an intervention being both less costly and more effective than the comparator.

### Data extraction

We extracted and reported data related to interventions, methods, and results, with multiple data points for studies reporting multiple interventions meeting inclusion criteria. Two reviewers (IR and DB) independently extracted data and assessed quality using a structured Excel spreadsheet. Differences between reviewers were reconciled by discussion, with recourse to a third reviewer (OC) if necessary.

### Study quality

We assessed quality of reporting using the Consolidated Health Economic Evaluation Reporting Standards (CHEERS)(Husereau et al., 2013), which is widely used in systematic reviews of economic evaluations. We followed a common scoring approach (Hope et al., 2017; Mangham-Jefferies et al., 2014) by awarding studies 1 point for each CHEERS item “fully met”, 0.5 for “partially met”, and 0 for “not met” or when insufficient information was reported (Supplementary Material C). The item was coded as missing if not applicable. We calculated a percentage score, giving all criteria equal weight, and excluding “not applicable” items from the denominator for that study. While recognising the arbitrary nature of cut-offs, we assigned illustrative labels to studies based on their percentage score, with studies scoring ≥75% labelled as “high” quality, those scoring 60-74% as “medium”, 45-59% as “low” and <45% as “very low”.

### Data synthesis

To compare study results, we converted “cost per outcome” metrics to US$ in 2024 prices (Turner et al., 2019). We first converted to local currency using World Bank (2025c) rates for the study year, then adjusted for inflation in the study country using World Bank (2025a) deflators, then converted to 2024 US$. Studies not specific to a country were adjusted using US inflation. Guidance recommends against meta-analysis in most systematic reviews of economic evaluations because costs are highly specific to settings and time (Mandrik et al., 2021; van Mastrigt et al., 2016). We therefore compare metrics in a narrative synthesis to qualitatively assess the overall strength of the evidence and of methods used. We draw conclusions based only on studies judged medium or high quality, though we describe methods and results of all included studies.

Country-specific cost-effectiveness thresholds should reflect national resource availability and capture health benefits forgone if resources were withdrawn from interventions already funded (Chi et al., 2020; Drake et al., 2023). If interventions above the threshold for a given country are funded, they may displace more health benefits than they generate. For studies that reported cost per disability-adjusted life year (DALY) averted, we compare to the supply-side (opportunity cost) thresholds for the respective country estimated by Ochalek et al. (2018), updated to 2024. We consider these to provide a “plausible” range. For the adjustment, we took the highest and lowest of the percentages of GDP per capita estimated by Ochalek et al. (2018) using four different methods (Bath et al., 2021). We then applied those two percentages to GDP per capita for 2024 using World Bank (2025b) data to generate a range for the 2024 supply-side threshold. We compare studies’ results from the provider perspective, where available, to that range. Given recent global health budget cuts, the lower end of the range is likely to be more appropriate. As a sensitivity analysis, we also compare results to a range for cost per quality-adjusted life year (QALY) gained estimated by Pichon-Riviere et al. (2022), further explained in Supplementary Material D. We do not make comparisons to 1 x GDP per capita or 3 x GDP per capita, because these old rules of thumb are now strongly discouraged as failing to reflect opportunity cost of health resources (Robinson et al., 2017).

We also categorised studies in several ways. First, we distinguish between empirical and modelled studies (Mandrik et al., 2021). Empirical studies collect primary data alongside and/or following actual implementation of an intervention, sometimes supplemented by secondary data. Modelled studies evaluate scenarios constructed exclusively from secondary data, rather than from a single occurrence of implementation that actually happened. Second, we distinguish between studies that incorporated modelling of adherence to handwashing over time after the intervention (using primary or secondary data) and those that did not.

## 3. Results

### Identification, screening, eligibility, and inclusion

Searches of electronic databases and websites yielded 2,610 results, with a further 10 identified from previous reviews and the study team (Figure 1). After removing duplicates, we screened titles and abstracts of 1,369 unique publications, and reviewed 45 full texts. Following full-text review, we included 15 studies. For the 30 studies excluded in full text review, the most common reasons were an ineligible intervention or only reporting a partial (costs only) economic evaluation (Figure 1, Supplementary Material E).

**Figure 1.**
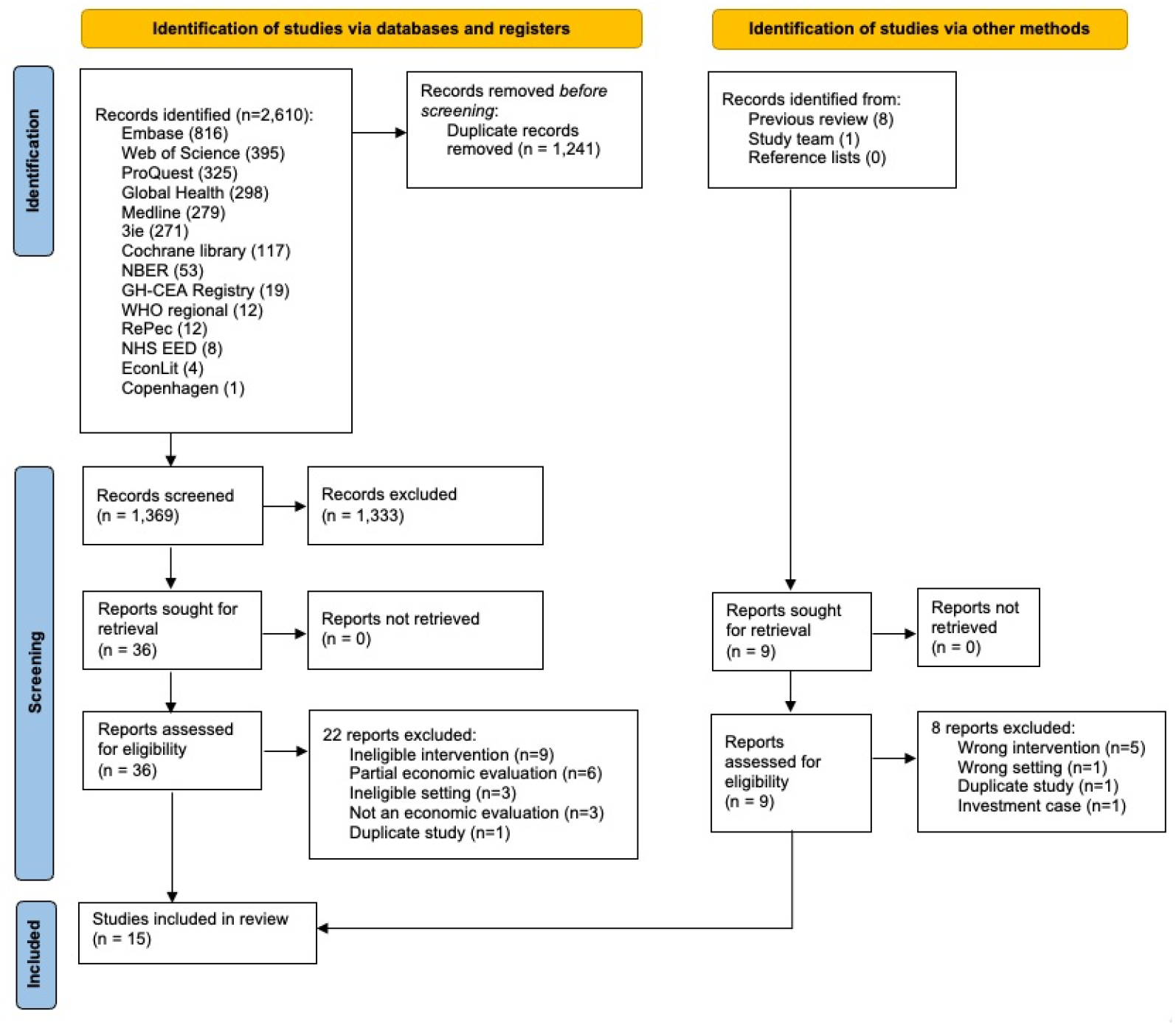
PRISMA flow diagram.

### Study settings and interventions

Of the 15 included studies, 8 were in specific low- and middle-income countries (L&MICs), 4 in generic L&MIC settings, and 3 in specific high-income countries (HICs)(Table 1). Most interventions assessed behaviour change in domestic settings (n=13), though 1 assessed a childcare setting (Azor-Martinez et al., 2021) and 1 university accommodation (Lachance, 2010). No interventions were in humanitarian settings.

**Table 1:** Study characteristics.

| Reference | Country & setting | Intervention | Outcome inference (if primary data) | Population (if primary data) | Health benefits estimated from primary data or assumed from literature |
| --- | --- | --- | --- | --- | --- |
| **Cost-effectiveness analyses** |  |  |  |  |  |
| Varley (1998) | Generic (urban) | **Hypothetical** - two eligible interventions modelled: (i) hygiene only (education on personal / food / domestic hygiene, excreta disposal and water storage); (ii) hygiene added to public tap installation | n/a (secondary) | n/a (secondary) | **Assumed** - from unpublished review of diarrhoea morbidity: (i) 10% reduction with hygiene only; (ii) 20% incremental reduction with hygiene added to existing public taps |
| Mascie-Taylor (1999) | Bangladesh (rural) | **Empirical** - hygiene promotion (after baseline albendazole) with monthly home visits, group discussions and school visits, focused on handwashing and other areas of hygiene | Before-after comparison without control, no year | 550 children aged 2-8 years | **Estimated within-study** - prevalence of roundworm, whipworm, and hookworm in stool by Kato-Katz method (weighted 28% reduction) |
| Borghi (2002) | Burkina Faso (urban) | **Empirical** - hygiene promotion with monthly home visits, health centre talks, radio programmes, etc., focused on handwashing and safe disposal of child stool | Before-after comparison without control, 1995-1998 | 289 mothers of children aged ≤36 months | **Assumed** - from mean of 6 diarrhoea morbidity studies: 42% reduction. Justified on basis of primary structured observation of handwashing after cleaning child's anus (13% before, 31% after) |
| Larsen (2003) | Generic (global) | **Hypothetical** - non-specific "hygiene" intervention but uses Varley (1998) cost data so assume similar (education on personal/food/domestic hygiene, excreta disposal and water storage) | n/a (secondary) | n/a (secondary) | **Assumed** - from discussion of various studies of diarrhoea, literacy, and mortality: 25-48% reduction in child mortality |
| Cairncross (2006) | Generic (national) | **Hypothetical** - handwashing promotion with unspecific content but costs derived from Varley (1998), Borghi (2002), and two other studies | n/a (secondary) | n/a (secondary) | **Assumed** - from review of diarrhoea morbidity (Curtis & Cairncross, 2003): 48% reduction |
| Hansen (2008) | Zimbabwe (national) | **Hypothetical** - "vertical programme" to promote unspecific "personal and domestic hygiene" | n/a (secondary) | n/a (secondary) | **Assumed** - from 3 reviews: exact outcome and reduction unclear |
| Lachance (2010) | USA (university) | **Empirical** - website promoting handwashing with soap, covering coughs/sneezes, and masking, alongside provision of hygiene kit (sanitiser, facemasks, thermometer) | n/a - RCT found no effect, so inferred outcomes from systematic review | n/a (secondary) | **Assumed** - from review of respiratory morbidity (Aiello et al., 2008): 21% reduction |
| Machdar (2013) | Ghana (urban) | **Hypothetical** - unspecific "hygiene" intervention but uses Borghi (2002) cost data so assume similar focused on handwashing (home visits, talks, radio), plus supply of chlorine for stored water disinfection | n/a (secondary) | n/a (secondary) | **Assumed** - uses reviews of handwashing (Sijbesma et al., 2009) and water quality to make unclear inference about 1-2 log-reduction in pathogen concentration. Converts to DALYs via Quantitative Microbial Risk Assessment |
| Sardar (2013) | Zimbabwe (national) | **Hypothetical** - unspecific promotion of hand hygiene with soap provision, alongside clean water distribution | n/a (secondary) | n/a (secondary) | **Assumed** - from unspecific source: appears 20% reduction in cholera morbidity (but unclear) |
| Siu (2021) | Gambia (rural) | **Empirical** – handwashing with soap and food hygiene promotion with home visits, community meetings, drama/songs | Cluster-randomised controlled trial, 2015-17 | Intervention: 377 mothers of children 6-36 months. Control: 370 mothers | **Estimated within-study** - 60% reduction in childhood diarrhoea at 6 months post-intervention, falling to 32% at 24 months (suggesting effects may have attenuated to zero eventually). |
| Azor-Martinez (2021) | Spain (childcare) | **Empirical** - soap arm included pre-intervention parents/staff workshop, with promotion of HWWS at 7 critical times, and informational brochure provided. Liquid soap dispensers installed in classrooms and supplies given for home use | Cluster-randomised controlled trial, 2013-14 | Intervention: 274 children aged 0-36 months. Control: 298 children | **Estimated within-study** - 6% reduction (not statistically significant at 5% level) in respiratory infection versus control. Nb. there was also a sanitiser arm which was more effective. |
| Beresniak (2023) | Italy (national) | **Hypothetical** - unspecific government promotion of handwashing, mask wearing, respiratory etiquette | n/a (secondary) | n/a (secondary) | **Assumed** - unspecified "panel of experts" estimated a 1–5% probability of achieving mortality reduction $\geq 40\%$, and the model sampled from that range with a uniform distribution |
| **Benefit-cost analyses** |  |  |  |  |  |
| Whittington (2012) | Generic (rural) | **Hypothetical** - unspecific handwashing promotion, but acknowledges diversity in scale/design and estimates cost ranges to reflect this | n/a (secondary) | n/a (secondary) | **Assumed** - from 2 reviews of diarrhoea morbidity: 45% reduction |
| Larsen (2016) | Bangladesh (rural) | **Hypothetical** - unspecific handwashing with soap promotion programs targeting mothers with young children | n/a (secondary) | n/a (secondary) | **Assumed** - from burden of disease study (Prüss-Ustün et al., 2014): 23% reduction in diarrhoea morbidity |
| Townsend (2017) | India/China (national) | **Hypothetical** - unspecific handwashing promotion, acknowledges diversity in design | n/a (secondary) | n/a (secondary) | **Assumed** - from review of diarrhoea morbidity (Freeman et al., 2014): 40% reduction |

HWWS = handwashing with soap, RCT = randomised controlled trial

Five studies promoted handwashing only, and 4 promoted broad or unspecific “hygiene” including handwashing (Table 1). The other 6 studies promoted handwashing alongside other specific behaviours, namely 2 with respiratory hygiene, 1 with food hygiene, 1 with child faeces disposal, 1 with water chlorination, and 1 with water supply. For 13 studies, the comparator was “no intervention” or “existing practice”, and 2 trial-based studies had active controls, namely thermometer provision (Lachance, 2010) and vegetable gardening (Siu et al., 2021)(Supplementary Material F).

No studies evaluated more than one handwashing with soap intervention. However, Varley et al. (1998) modelled the same handwashing with soap intervention in two different contexts, one with unimproved water (e.g. unprotected wells) and one with public taps. One study compared two hand hygiene interventions to a control, but one intervention used sanitiser so did not meet inclusion criteria (Azor-Martinez et al., 2021).

### Study design

Of the 15 studies, 4 used BCA, 11 used CEA, and none used both (Table 1). Only 5 studies were empirical (some primary data alongside an actual intervention), and most (n=10) were modelled (wholly secondary data evaluating a hypothetical intervention). All n=10 modelled studies assumed health benefits from the literature, as did 2 of the empirical studies, and only 3 empirical studies estimated health effects directly (Table 1). Most studies (n=11) valued reductions in diarrhoea risk only (all in L&MICs), 3 valued respiratory infections only (all in HICs), and none valued both. The remaining study valued helminth infections only. Studies assuming impact on diarrhoea from the literature employed a wide range of risk reductions (10% to 48%).

Cost per DALY averted was reported in 8 studies, cost per QALY gained in 1, and a benefit-cost ratio in 4 (Table 2). Two studies reported only cost per case averted, and 2 reported only intervention-specific measures that cannot easily be compared across studies (e.g. cost per weighted percentage point reduction in three helminths).

**Table 2:** Study results.

| Reference | Intervention type | Measures | Key result (2024 US\$) and perspective | Results (original currency and year) | Handwashing adoption and time horizon | Study quality |
| --- | --- | --- | --- | --- | --- | --- |
| **Cost-effectiveness analyses** |  |  |  |  |  |  |
| Varley (1998) | broad hygiene added to unimproved water | Cost per DALY / case / death averted | \$81 / DALY averted (provider perspective) - see Figure 2 | hygiene alongside unimproved water: US\$ 44/DALY averted; \$6/case averted; 1,520/death averted (1996) | **Adoption not measured or modelled** - assumes sustained behaviour change (commensurate with assumed disease reduction) over 80-year horizon | medium |
| | broad hygiene added to public taps | | \$37 / DALY averted (provider perspective) - see Figure 2 | hygiene alongside public taps: US\$ 20/DALY averted; \$3/case averted; \$689/death averted (1996) | | |
| Mascie-Taylor (1999) | broad or unspecific hygiene | Cost per weighted % reduction in prevalence of 3 worms | \$1,631 / % reduction (appears provider perspective) | 36,313 BDT (1997 assumed) | **Adoption not measured or modelled** - applies observed disease reduction over 18-month horizon | low |
| Borghi (2002) | handwashing and child faeces disposal | Cost per DALY / case / death averted | \$96 / DALY averted (provider perspective, societal also reported) - see Figure 2 | US\$ 90 / DALY averted (2012 prices) as converted by Horton et al. (2017). Original estimates in Borghi et al. (2002) from provider perspective were \$34 / case averted (1999 prices) and \$2,792 / death averted. | **Adoption measured** - structured observations before/after intervention aren't part of CEA model (assumes disease reduction from literature over 3-year horizon) | high |
| Larsen (2003) | broad or unspecific hygiene | Cost per DALY / death averted | \$28 / DALY averted (appears societal perspective) | US\$ 15 / DALY averted, US\$ 500 / death averted (1996 assumed as uses Varley costs) | **Adoption not measured or modelled** - effectively assumes sustained behaviour change (commensurate with assumed disease reduction) over an unclear time horizon | very low |
| Cairncross (2006) | handwashing | Cost per DALY averted | \$6 / DALY averted (appears provider perspective) | US\$ 3.35 (2000 assumed) | **Adoption not measured or modelled** - effectively assumes sustained behaviour change (commensurate with assumed disease reduction) over an unclear time horizon, but acknowledges promotion requires repetition every 5 years | low |
| Hansen (2008) | broad or unspecific hygiene | Cost per DALY averted | \$107 / DALY averted (provider perspective) | US\$ 59 (1997) | **Adoption not measured or modelled** - effectively assumes sustained behaviour change (commensurate with assumed disease reduction) over an unclear time horizon | low |
| Lachance (2010) | hand washing and respiratory hygiene | Cost per QALY gained | \$82,967 / QALY gained (societal perspective) | US\$ 58,310 (2008) | **Adoption not measured or modelled** - applies disease reduction from literature over 18-month horizon | high |
| Machdar (2013) | handwashing with water chlorination | Cost per DALY averted | \$7 / DALY averted (appears provider perspective) | US\$ 6 (2011 assumed) | **Adoption not measured or modelled** - effectively assumes sustained behaviour change (commensurate with assumed disease reduction) over an unclear time horizon | very low |
| Sardar (2013) | handwashing with water supply | Cost per cholera case averted | \$138 / case averted (appears provider perspective ) | US\$ 101 (median across 9 regions, 2011 assumed) | **Adoption not measured or modelled** - effectively assumes sustained behaviour change (commensurate with assumed disease reduction) over an unclear time horizon | low |
| Siu (2021) | handwashing and food hygiene | Cost per DALY / case /death averted | \$994 / DALY averted (societal perspective) - see Figure 2 | US\$ 814 / DALY; US\$ 30,786 / death (2017) | **Adoption measured** - structured observations of handwashing at 0, 6, 32 months (reported in trial paper), but aren't part of CEA model, which assumes observed disease reduction sustained over 4-year horizon | high |
| Azor-Martinez (2021) | handwashing | Cost per respiratory case averted | Dominant, saves \$30 / case (societal, provider also reported) | Dominant, with cost saving of EUR 27 (2018) per case averted versus control (95% CI: -115 to 61), with 72% probability of dominance (more effective, less costly) in PSA | **Adoption not measured or modelled** - applies observed disease reduction over 8-month horizon | high |
| Beresniak (2023) | hand washing and respiratory hygiene | Average cost per "success" (achieving mortality reduction ≥40%) | \$323 million / success (provider / system perspective) | 254 million euros (2017) | **Adoption not measured or modelled** - applies assumed disease reduction over 9-month horizon | low |
| **Benefit-cost analyses** |  |  |  |  |  |  |
| Whittington (2012) | handwashing | Benefit-cost ratio | 2.1 (societal perspective) | 2.1 in "medium" adoption/adherence scenario (2010 prices assumed) | **Adoption modelled** - applies adoption/adherence estimates from literature to inform assumptions about disease reductions over 18-month time horizon | high |
| Larsen (2016) | handwashing | Benefit-cost ratio | 1.08 (societal perspective) | 1.08 (2014 prices) | **Adoption not measured or modelled** - applies disease reduction from literature over 3-year horizon | low |
| Townsend (2017) | handwashing | Benefit-cost ratio | 92 India<br>35 China (societal perspective) | 92 India, 35 China (2015 prices assumed) | **Adoption not measured or modelled** - applies disease reduction from literature over 1-year horizon | very low |

DALY = disability-adjusted life year, QALY = quality-adjusted life year, PSA = probabilistic sensitivity analysis

A provider perspective only was taken in 8 studies (explicitly or implicitly), while 5 took a societal perspective only, with 2 reporting results from both perspectives (Azor-Martinez et al., 2021; Borghi et al., 2002) (Supplementary Material F). The time horizon of models was unclear in five studies and ranged from 8 months to 80 years in the remaining 10 studies (Table 2), indicating a wide variety of approaches to modelling disease and behaviour. Amongst the 10 studies with clear horizons, the median was 1.5 years. Simple deterministic sensitivity analysis (varying one parameter at a time) was reported in 7 studies, 3 included a probabilistic sensitivity analysis (varying many parameters simultaneously) and 5 reported no sensitivity analysis (Supplementary Material F).

Of the 5 empirical studies, only 2 measured handwashing behaviour change, both using structured observations. Both saw modest improvements in observed handwashing with soap at critical times, from 13% to 31% in Borghi et al. (2002) and 11% to 19% in Siu et al. (2021)(Table 2), though these were not explicitly part of the analysis and health benefits were estimated directly. The remaining 3 empirical studies did not measure handwashing behaviour.

Of the 10 modelled studies, only 1 incorporated estimates of behavioural adoption into the model (Whittington et al., 2012). Specifically, the authors modelled the proportion of targeted households who adopt handwashing with soap (median 40% based on literature review). They also modelled the proportion of adopting households who adhere to the behaviour for the rest of the 1.5 year time horizon (median 50%). The achievement of benefits is then determined by these parameters. Amongst the remaining 9 modelled studies which but did not model behavioural adoption, 6 assumed that behaviour change was sustained for more than 3 years without attenuation.

### Study quality

Five studies were scored as high quality, 1 as medium, 6 as low, and 3 as very low. Percentage scores ranged 36%-90%, with median 55%. Empirical studies tended to score more highly (median 82%) than modelled studies (median 52%). Distributions of scores per CHEERS item are presented in Supplementary Material G, alongside a heat map for item scores, and a plot of study quality over time (which does not appear to show an improvement).

Across CHEERS items, studies were most likely to adequately describe which outcomes were used as the measure(s) of benefit (93% fully met, 0% not met) and least likely to describe the effects of uncertainty (13% fully met, 60% not met). Authors tended to give more attention to effects than to costs, with only 27% fully meeting the CHEERS items for resources/costs. Only 5 studies collected primary cost data from clearly-reported sources, that is, all 5 empirical studies. Amongst the 10 studies using secondary cost data, only 3 scored “fully met” for the costing CHEERS item (Supplementary Material G). Reasons for lower scores included limited information being provided on cost data sources and assumptions, being unclear on what activities/interventions were assumed to be costed, and using secondary cost data without explanation.

### Cost-effectiveness results

Of the 6 medium- or high-quality cost-effectiveness studies, 3 reported cost per DALY averted (Figure 2) as an incremental cost-effectiveness ratio (ICER). Borghi et al. (2002) report an empirical study of an intervention promoting handwashing and child faeces disposal to mothers in urban Burkina Faso, using secondary data for health impact (Table 1). The intervention was relatively large-scale, being delivered across all administrative areas of a town of 300,000 people over 3 years. Horton et al. (2017) use Borghi et al.’s result for cost per death averted (provider perspective) to estimate US$ 96 per DALY averted (2024 prices), by assuming that a death in early childhood is equivalent to 32 discounted DALYs (Solberg et al., 2020). Both Borghi and Horton conclude that the intervention was cost-effective. This appears justified in comparison to the range of plausible supply-side thresholds for Burkina Faso in 2024 (Figure 2). In a sensitivity analysis using the Pichon-Riviere et al. thresholds (Supplementary Material D), the intervention would still be cost-effective for most of the range.

**Figure 2.**
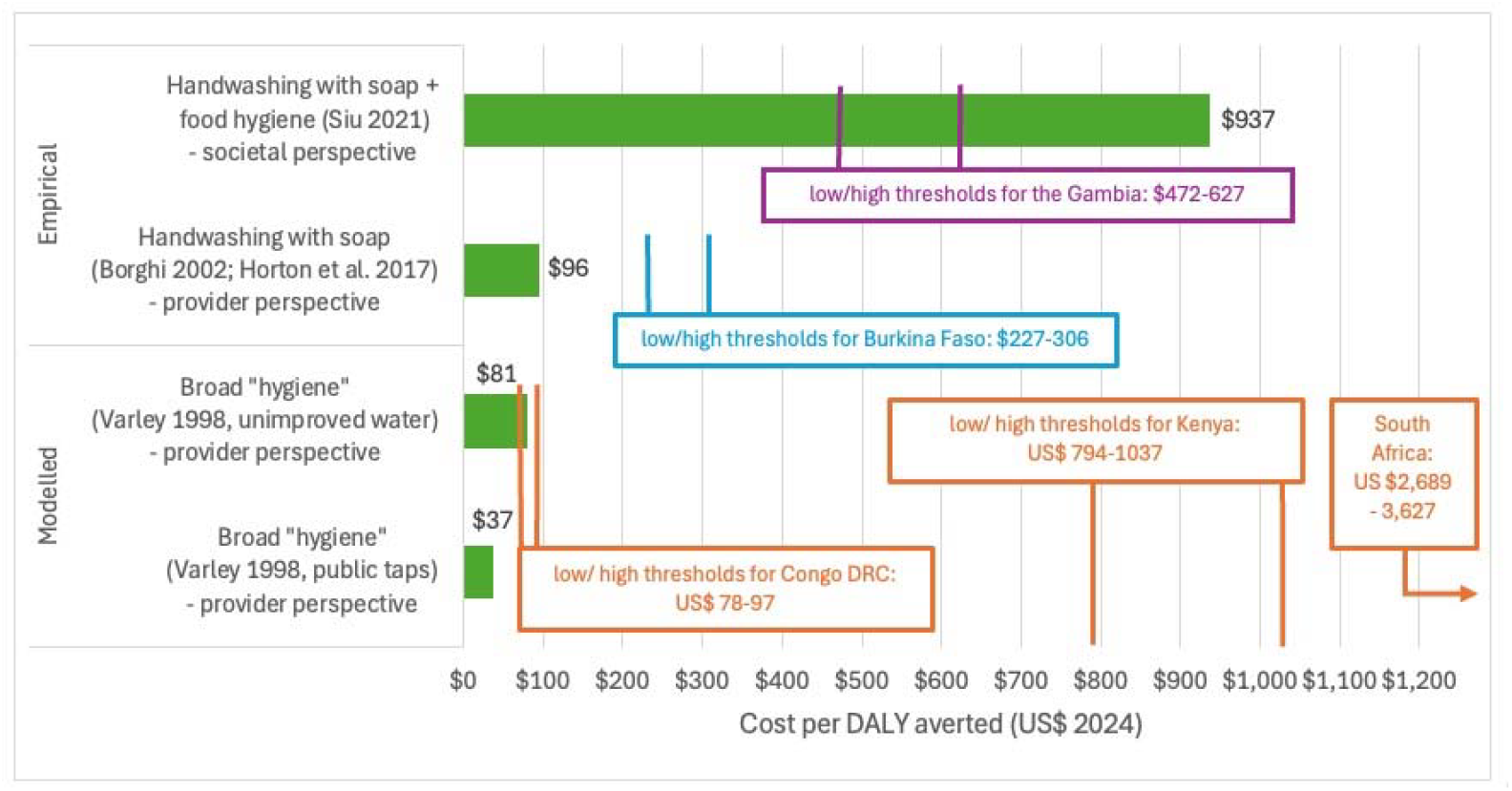
Incremental cost per DALY averted for handwashing interventions in high- or medium-quality studies. Data are presented in US$ 2024, and cost-effectiveness thresholds are from Ochalek et al. (2018). For visualisation of results compared to alternative thresholds from Pichon-Riviere et al. (2023), see Supplementary Material D. The Borghi et al. result for DALYs was estimated by Horton et al. (2017), see Supplementary Material D. Studies did not present confidence intervals for incremental cost-effectiveness ratios.

Siu et al. (2021) report an empirical study alongside a randomised controlled trial of an intervention promoting handwashing and food hygiene to mothers in rural Gambia (Table 1). The intervention was relatively small-scale, being delivered across 30 villages (population around 10,000). The authors estimate US$ 937 per DALY averted (societal perspective, 2024 prices) and conclude that the intervention was cost-effective in relation to a threshold of 3 x GDP per capita but not cost-effective in relation to a supply-side threshold. Comparisons to supply-side thresholds would ideally be taken from the provider perspective (rather than societal). Nonetheless, this intervention seems quite unlikely to be cost-effective in comparison to plausible supply-side thresholds for the Gambia in 2024 (Figure 2), and very unlikely to cost-effective using the Pichon-Riviere et al. thresholds (Supplementary Material D). Both the Siu et al. and Borghi et al. studies were undertaken after formative research and with researcher-supported delivery, rather than being implemented by the health system in general.

Varley et al. (1998) report a modelled study of the same broad-based hygiene intervention in two hypothetical water supply contexts, one costing US$ 37 per DALY averted and the other $81 per DALY averted in 2024 prices (Figure 2). No country is modelled but rather a large city in a generic setting characterised by “slums” with few formal public services. The authors conclude that the intervention is cost-effective. Given the lack of a specific country, we present results (Figure 2) against some plausible thresholds for three African countries at low, medium and high points in the distribution of GDP per capita. In all three cases, the intervention would be likely to be cost-effective.

There were two medium- or high-quality studies in high-income countries. The first evaluated a handwashing intervention in a Spanish childcare setting (Azor-Martinez et al., 2021). It was cost-saving compared to control from the societal perspective, and therefore highly cost-effective. The second evaluated a handwashing and respiratory hygiene intervention in USA student accommodation, and cost $82,967 per QALY gained from societal perspective. This is cost-effective compared to the $100,000 per QALY threshold predominantly applied in the USA (Neumann & Kim, 2023).

Of the 9 low- and very-low-quality studies, 4 reported a cost per DALY averted, with ICERs ranging from US$ 5 to 101 per DALY averted. This would be cost-effective in most L&MICs – see the threshold for the Democratic Republic of the Congo in Figure 2 which is the lowest amongst Ochalek et al. (2018) estimates, and about 10 times lower than the Kenya threshold.

### Benefit-cost results

The only medium- or high-quality benefit-cost analysis was a modelled study of a generic community-based intervention promoting handwashing with soap (Whittington et al., 2012). Assuming “medium” levels of handwashing adoption and adherence, the study estimated a mean benefit-cost ratio (BCR) of 2.1, meaning each US$ 1 invested in the intervention returned outcomes for society valued at $2.1. One-way sensitivity analyses indicated that the intervention would achieve BCRs greater than 1 with different proportions of the population adopting the behaviour (BCR range 1.7 – 2.3) and adhering to it over time (1.2 – 2.6)(Figure 3). However, when adoption and adherence were both low (20%), the mean BCR was 0.9, indicating costs higher than benefits. Amongst the 2 low- and very-low-quality benefit-cost analyses, BCRs ranged from 1 to 92.

**Figure 3.**
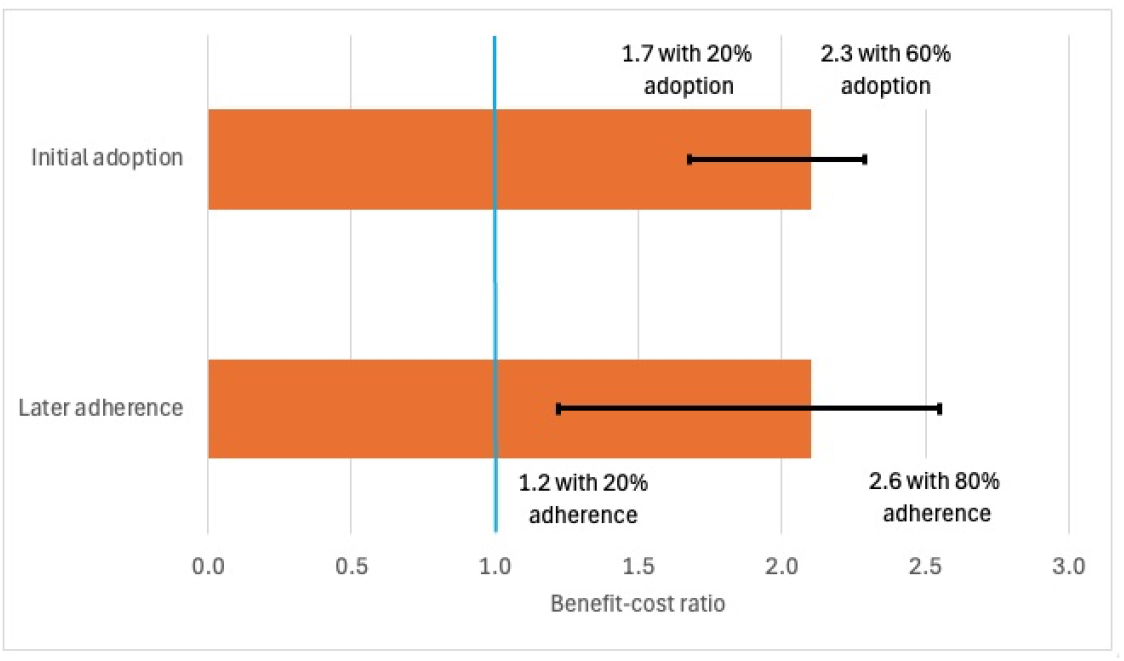
Benefit-cost ratios under one-way sensitivity analyses for initial handwashing adoption and later adherence (Whittington et al., 2012) Length of the horizontal bars is the benefit-cost ratio (2.1) when all parameters are at median values. Error bars show deterministic sensitivity analyses with other parameters remaining at median values. In the upper bar adherence is set at 50% when adoption is being varied, and the lower bar adoption set at 40% when adherence is being varied. The blue line shows break-even where benefits equal costs.

## 4. Discussion

Our findings show that promotion of handwashing with soap in domestic, school and childcare settings is very likely to be cost-effective for interventions that successfully increase and sustain adoption of handwashing behaviours. Interventions achieving handwashing adoption are also likely to have benefits greater than costs. However, evidence is scarce and of wide-ranging quality. Maximising adoption requires careful planning and delivery, plus tailoring to behavioural determinants in target populations (Caruso et al., 2025; Prasad et al., 2025; White et al., 2020). The wide range of results for cost per DALY averted (Figure 2) is not unusual in systematic reviews of economic evaluations; what is important is comparison of results to country-specific cost-effectiveness thresholds. Of the 3 estimates, the 2 taking the provider perspective were cost-effective compared to plausible supply-side thresholds for the respective country (Figure 2). The 2 empirical studies in Figure 2 were undertaken with researcher-supported delivery and after extensive formative research. Though the Burkina Faso study was already at a reasonable scale (300,000 population) effectiveness may experience a “scale-up penalty” (Chase & Do, 2012; Galiani et al., 2015; McCrabb et al., 2019), though this could be offset by lower unit costs (Ross et al., 2021). Reasons for thinking this literature understates cost-effectiveness of handwashing promotion include that none of the studies in L&MICs which valued diarrhoea reductions also valued reductions in respiratory infections, despite the substantial attributable burden preventable by handwashing with soap (Ross et al., 2023; Wolf et al., 2023). Second, no study empirically measured quality of life benefits beyond disease (e.g. feelings of pride or cleanliness)(Whittington et al., 2012). Handwashing promotion can be cost-effective in higher-income settings too; interventions in a Spanish childcare setting and USA student accommodation were cost-saving and cost-effective, respectively. In healthcare settings, interventions to improve hand hygiene tend to be cost-effective or cost-saving (Rice et al., 2023). Though the evidence for domestic and other settings assessed here is less clear cut, promoting handwashing with soap can in many cases be as cost-effective for child health as pneumococcus/rotavirus vaccinations and oral rehydration therapy (Horton, 2018).

Strengths of our review include that we assessed studies’ quality of reporting. Ratings using CHEERS strongly correlate with our opinion of the rigour of the studies, so we believe the better-reported studies indeed provide a more reliable basis for conclusions. Also, we extracted and reported many aspects of study design, which could help future researchers avoid repeating prior weaknesses. Limitations of our review include that we focused on handwashing with soap, to the exclusion of sanitiser-based interventions, which may play an important role in some settings. While having used CHEERS is a strength, it does measure quality of reporting rather than actual study or model quality. Future systematic reviews might consider using the Criteria for Health Economic Quality Evaluation (CHEQUE), which was designed to differentiate methods from reporting, and to account for the relative importance of quality attributes (Kim et al., 2023). Finally, there remains much debate on which cost-effectiveness thresholds are appropriate for different countries and different decision-makers (Chi et al., 2020; Drake et al., 2023). We applied two sets of supply-side thresholds (Ochalek et al. (2018) and Pichon-Riviere et al. (2022)), but in the context of recent reductions in global health funding, these may be too high in the short-term. This uncertainty over appropriate thresholds means our conclusions about cost-effectiveness cannot be as clear-cut as we would like. For example, if the opportunity cost of health resources in Burkina Faso is at the lower bound of Pichon-Riviere et al. estimates, then even the intervention evaluated by Borghi et al. (2002) may not be cost-effective (Supplementary Material D).

Our systematic review addressed many limitations of the only previous review in this area (Hutton & Chase, 2016), and we identified twice as many studies. We also diagnose relatively low study quality, which does not appear to have improved over time (Supplementary Material G). Specific literature gaps to be addressed include, first, that more empirical studies with primary data collection are required. In many areas of health, an important role in the literature is played by trial-based economic evaluations measuring outcomes and costs in the same population (Ramsey et al., 2015), but such studies are under-represented in the handwashing literature. Only 2 of the 12 studies in L&MIC settings applied both primary cost and primary outcome data (Mascie-Taylor et al., 1999; Siu et al., 2021). Second, no diarrhoea-oriented study in a L&MIC also valued reductions in respiratory diseases. Third, no study compared two or more interventions promoting handwashing with soap (in addition to existing practice /doing nothing) which would better reflect real-world choices faced by decision-makers, e.g. between interventions of different intensities (Pickering et al., 2019). Fourth, only one study directly modelled behaviour parameters (Whittington et al., 2012). The extent to which handwashing behaviours exist pre-intervention, are adopted post-intervention, and are sustained over time, are crucial factors in whether economic benefits are achieved. If health benefits are directly measured, then not incorporating adoption into the model is justifiable. Even then, however, behaviour should be measured or assumptions about adoption and adherence over time should be clarified. If health benefits are not directly measured, then it is even more important to undertake measurement or modelling of behaviours, or at the very least make assumptions clear. Amongst the 9 modelled studies which did not incorporate adoption variables, 6 assumed that behaviour change was sustained for more than 3 years without attenuation. Such assumptions require strongly-evidenced justification, since various studies show both handwashing behaviour and health effects tailing off sooner (De Buck et al., 2017; Luby et al., 2009; Manaseki-Holland et al., 2021).

Our results can support improvement in methodological and reporting quality of cost-effectiveness and benefit-cost analyses of handwashing promotion. They can also contribute to debates around the prioritisation of hygiene, and of intervention options within hygiene, in the context of upcoming WHO guidelines and the Lancet Commission on WASH (Amebelu et al., 2021). Transferring economic evaluation results across settings can be challenging due to heterogeneity in various factors (e.g. epidemiology, macroeconomy, and health system capacity). Just because an intervention is cost-effective in one health system does not mean it will be in another, because of how the above factors influence incremental costs, outcomes and thresholds. For example, see the very different thresholds illustrated for three countries in relation to Varley et al. (1998) in Figure 2.

We have several recommendations for policy, practice, and research. First, interventions should be carefully planned to maximise and sustain handwashing adoption, while addressing behavioural determinants in the target population. Second, to enable clear decisions about priorities, governments should either have a dedicated budget for hygiene or be able to identify hygiene expenditures across ministries. In a WHO-led exercise on resources for water, sanitation and hygiene, only 17% of 109 countries could report their hygiene budget and/or expenditures (WHO, 2022). Third, research is needed to address the literature gaps identified above, especially more empirical studies which value both respiratory and diarrhoea diseases, compare multiple feasible promotion interventions, and adequately measure and/or model adoption of handwashing over time.

## Data Availability

All relevant data are within the manuscript and its Supporting Information files.

## Contributors

Ian Ross: conceptualisation, literature search, data accessed and verified, data analysis, data interpretation, writing - original draft, writing - review & editing, decision to submit

David Bath: conceptualisation, data accessed and verified, data analysis, data interpretation, writing - review & editing

Joseph Wells: data accessed and verified, data analysis, writing - review & editing

Robert Dreibelbis: conceptualisation, writing - review & editing

Regina Ejemot-Nwadiaro: conceptualisation, writing - review & editing

Joanna Esteves Mills: writing - review & editing

Giulia Greco: conceptualisation, writing - review & editing

Catherine Pitt: conceptualisation, writing - review & editing

Oliver Cumming: conceptualisation, data accessed and verified, data interpretation, writing - review & editing, supervision, decision to submit.

## Acknowledgements

We thank authors of included studies who responded to requests for more information. Ian Ross acknowledges the support of a fellowship from the Reckitt

Global Hygiene Institute in the period when the paper was drafted.

## Tables and Figures

### Figures

Figure 1. Study profile

Figure 2. Incremental cost per DALY averted for handwashing interventions in high- or medium-quality studies

Figure 3. Benefit-cost ratios under one-way sensitivity analyses for initial handwashing adoption and later adherence (Whittington et al., 2012)

