## Supplementary Materials for "Cost-effectiveness and benefit-cost analyses of promoting handwashing with soap: a systematic review"

### A. Search strategy

As applied on OVID-SP interface (Medline, EMBASE, Global Health, Econlit)

|  | | HANDWASHING WITH SOAP |
| --- | --- | --- |
| 1 | | (handwash* or (hand* adj3 (wash* or hygiene))).mp |
| 2 | | (hand* adj3 soap*).mp |
| 3 | | ((hygiene or handwash*) adj3 (promot* or educat* or behavio*)).mp |
| 4 | | 1 or 2 or 3 |
|  | | ECONOMIC EVALUATIONS |
|  | 5 | (cost-benefit or benefit-cost or cost-effectiv* or cost-utility or (econ* adj1 eval*) or (cost adj2 efficien*) or value for money or value-for-money).mp |
|  | | COMBINING ALL TERMS |
| 6 | | 4 and 5 |
|  | | LIMITS |
|  | 7 | limit 6 to yr="1980 -Current" |
| 8 | | Limit 7 to (humans) |

As applied on Web of Science

|  | HANDWASHING WITH SOAP |
| --- | --- |
| # 1 | TS=(handwash* or (hand* NEAR/3 (wash* or hygiene) )) |
| # 2 | TS=(hand* NEAR/3 soap*) |
| # 3 | TS= ((hygiene or handwash*) NEAR/3 (promot* or educat* or behavio*) ) |
| # 4 | #3 OR #2 OR #1 |
|  | ECONOMIC EVALUATIONS |
| # 5 | TS= (cost-benefit or benefit-cost or cost-effectiv* or cost-utility or (econ* NEAR/1 eval*) or (cost NEAR/2 efficien*) or value for money or value-for-money) |
|  | COMBINING ALL TERMS |
| # 6 | #4 AND #6 |

We also searched the below databases and websites, adapting the above-listed search terms to their interfaces:

- Cochrane library,
- International Bibliography of the Social Sciences,
- Global Health Cost Effectiveness Analysis Registry,
- NHS Economic Evaluation Database
- International bibliography of the social sciences
- National Bureau of Economic Research
- International Initiative for Impact Evaluation
- Research Papers in Economics,
- WHO Index Medicus (all regions)
- Copenhagen Consensus Centre

### B. Categorisation of hygiene interventions

We included studies of combined interventions if they reported effect estimates separately for the handwashing component or clearly had handwashing as a “major” component, that is, any of the three categories in Table B-1. This follows the same categorisation as Ross et al. (2023)

**Table B‑1: Categories of the extent of combination of hygiene interventions**

|  | **Descriptor** |
| --- | --- |
| **1. Handwashing only** | Interventions focused on handwashing alone (or alongside other body washing, e.g. bathing, or face-washing) |
| **2. Handwashing majority (≥50% of messages)** | Interventions focused on handwashing alongside other components, but where the handwashing component was the behavioural target in ≥50% of intervention messages. No distinction is made regarding what handwashing is combined with (e.g. household water treatment, food hygiene). |
| **3. Handwashing minority (~25-50% of messages)** | Interventions where handwashing is a major part of a broad intervention, but is the behavioural target in <50% of intervention messages. To be included, the CEA/BCA itself (rather than a referenced paper, e.g. process evaluation), must indicate that handwashing is part of the intervention anywhere in the methods section. In cases where “hygiene” is mentioned rather than handwashing, but references and other details (e.g. soap) imply handwashing, this is interpreted as handwashing. |

### D. Scoring for CHEERS

This table below sets out the protocol we developed for scoring each CHEERS reporting item.

**Table C‑1: Categories of the extent of combination of hygiene interventions**

EE = economic evaluation, CEA = cost-effectiveness analysis, BCA = benefit-cost analysis, DALY = disability-adjusted life year

| **Area** | **Reporting item (from Husereau et al., 2013)** | | **Interpretation for scoring in this study (authors)** | | |
| --- | --- | --- | --- | --- | --- |
| **Framing** | **1. Title** | Identifies the study as an EE or uses specific terms such as CEA, and describes the interventions compared. | 1 - includes, CEA, BCA, EE, or similar, in the title | 0.5 - implication of EE but not with clear terms | 0 - nothing implies EE |
|  | **2. Abstract** | Structured summary of objectives, perspective, setting, methods, results (including base case & uncertainty) | 1 - abstract contains most of these | 0.5 - abstract provides some of above, but missing some of the most important (intervention, methods, results) | 0 - abstract provides very few of these |
|  | **3. Background and objectives** | Provides broader context for the study. Presents the study question and its relevance for policy or practice decisions. | 1 - study aim/question presented with relevance for decisions | 0.5 - EITHER clear aim but limited context OR good relevance re decisions, but unclear aim or question regarding EE | 0 - BOTH aim/question AND relevance for decisions unclear |
| **Population, setting** | **4. Target population and subgroups** | Describes characteristics of the base case population and subgroups analysed, including why they were chosen. | 1 - study population sufficiently described, including any assumptions | 0.5 - study population described, but too briefly in respect of policy-relevant characteristics | 0 - study population unclear |
|  | **5. Setting and location** | States relevant aspects of the system(s) in which the decision(s) need(s) to be made. | 1 - characteristics of decision context described | 0.5 - decision context described but insufficiently | 0 - decision context appears not to have been considered |
| **Key methods decisions** | **6. Study perspective** | Describes the perspective of the study and relate this to the costs being evaluated. | 1 - perspective stated clearly and interpreted | 0.5 - perspective vaguely stated, but sufficient detail such that it can easily be discerned | 0 - perspective unclear |
|  | **7. Comparators** | Describes the interventions or strategies being compared and state why they were chosen. | 1 - intervention and comparator both well-described | 0.5 - limited detail on comparator, but doesn’t substantially harm interpretation | 0 - lack of detail on comparator limits interpretation |
|  | **8. Time horizon** | States the time horizon(s) over which costs and consequences are being evaluated and say why appropriate. | 1 - time horizon stated and explained | 0.5 - time horizon stated without context, or clearly implied | 0 - time horizon unclear, which limits interpretation |
|  | **9. Discount rate** | Reports the choice of discount rate(s) used for costs and outcomes and say why appropriate. | 1 - rate reported and justified | 0.5 - rate reported but unclear how applied or not justified / referenced | 0 - no discounting, or unclear how applied |
| **Outcomes and costs** | **10. Choice of outcomes** | Describes what outcomes were used as the measure(s) of benefit in the evaluation and their relevance for the type of analysis performed. | 1 - outcomes / benefits clear | 0.5 - lack of clarity on some but not all benefits | 0 - outcomes / benefits very unclear |
|  | **11. Measurement of effectiveness** | *(i) Single study:* Describes features of design and why sufficient, (ii) *Synthesis-based*: describes identification & synthesis of included studies | 1 - source of effects data explained, discussed and justified | 0.5 - source and important assumptions clear, but not discussed / justified | 0 - source or important assumptions unclear |
|  | **12. Measurement & valuation of pref.-based outcomes** | If applicable, describe the population and methods used to elicit preferences for outcomes. | 1 - if DALYs used, source of weights referenced / discussed | 0.5 - slightly unclear on weights | 0 - very unclear on weights |
|  | **13. Estimating resources and costs** | Describes approaches used to estimate and value resource use, and any adjustments made to approximate to opportunity costs. | 1 - Approach to costing and sources of data are clear, with quality discussed | 0.5 - some information on costing missing, OR quality of data sources unclear / not discussed | 0 - much information / discussion of costing data sources missing |
|  | **14. Currency, price date, and conversion** | Reports the dates of the estimated resource quantities and unit costs, and methods for adjusting to the year/currency of analysis. | 1 - currency dates and conversions clear | 0.5 - minor issues of clarity | 0 - currency dates OR conversions not reported |
| **Modelling** | **15. Choice of model** | Describes and give reasons for the specific type of decision-analytical model used. | 1 - model is clearly described | 0.5 - model description has some limitations | 0 - model unclear |
|  | **16. Assumptions** | Describes all structural or other assumptions underpinning the decision-analytical model. | 1 - model assumptions clear, allowing reproducibility | 0.5 - most model assumptions clear, but not reproducible | 0 - model assumptions unclear |
|  | **17. Analytical methods** | Describes all analytical methods supporting the evaluation (e.g. missing or censored data; population heterogeneity) | 1 - explains important steps, e.g. how cost categories / types summed, dealt with missing data / outliers | 0.5 - partially explained | 0 - poorly explained |
|  | **18. Study parameters** | Report the values, ranges, references (and, if used, probability distributions) for all parameters. | 1 - values, ranges and references for input parameters reported | 0.5 - some model inputs described, but not values, ranges, references | 0 - input parameters poorly described |
| **Results and sensitivity** | **19. Incremental costs and outcomes** | Reports mean values for the main categories of estimated costs and outcomes, as well as mean differences between comparator groups | 1 - mean values for cost categories / outcomes reported | 0.5 - costs/outcomes only reported in aggregate, not disaggregated by category | 0 - very unclear reporting of costs / outcomes |
|  | **20. Characterising uncertainty** | Describes the effects of sampling and/or model uncertainty, with impact of methodological assumptions (e.g. discount rate) | 1 - parameter and structural uncertainty adequately characterised | 0.5 - only parameter OR structural uncertainty adequately characterised | 0 - limited characterisation of sources of uncertainty |
|  | **21. Characterising heterogeneity** | If applicable, reports differences in costs, outcomes, or cost-effectiveness that can be explained by variations between subgroups. | 1 - if sub-group analysis conducted, it is clear | 0.5 - if sub-group analysis conducted, it is fairly clear | 0 - if sub-group analysis conducted, it is unclear |
| **Other** | **22. Findings, limitations, generalisability** | Summarises key study findings and limitations, describing how results support the conclusions, and their generalisability. | 1 - findings / conclusions clearly linked and limitations discussed | 0.5 - conclusions unclear or not linked to findings, OR limitations not discussed | 0 - conclusions and limitations unclear |
|  | **23. Source of funding** | Describes how the study was funded and the role of the funder in the identification, design, conduct, and reporting of the analysis. | 1 - funder noted | n/a | . - funder unclear (not a norm) |
|  | **24. Conflicts of interest** | Describes any potential for conflict of interest of study contributors in accordance with journal policy. | 1 - COI adequately described | n/a | . - COI not described (not a norm) |

### D. Cost-effectiveness thresholds

Figure D‑1: Incremental cost per DALY averted for handwashing interventions in high- or medium-quality studies, using Pichon-Riviere cost-effectiveness thresholds for QALYs.

Data are presented in US$ 2024, and cost-effectiveness thresholds from those for quality-adjusted life years (QALYs) in Pichon-Riviere et al. (2023). While in theory gaining 1 QALY is equivalent to averting 1 DALY, these measures do have important distinctions in practice (Feng et al., 2020). The Ochalek thresholds have been more widely-used for DALY comparisons, which is why we do so in our study.

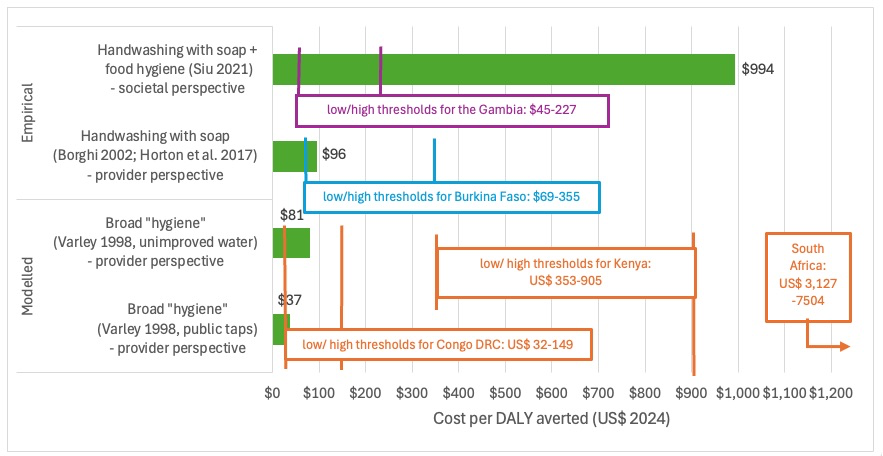

Table 3: Estimated thresholds in 2024 US$ based on percentages in two different studies applied to GDP per capita

|  | Ochalek et al. (2018) | | Pichon-Riviere et al. (2023) | |
| --- | --- | --- | --- | --- |
|  | low | high | low | high |
| Burkina Faso | 227 | 306 | 69 | 355 |
| The Gambia | 472 | 627 | 45 | 227 |
| Congo DRC | 78 | 97 | 32 | 149 |
| South Africa | 2689 | 3627 | 3127 | 7504 |
| Kenya | 794 | 1037 | 353 | 905 |

### E. Studies excluded at full-text review

**Table E‑1: Studies excluded at full-text review with reasons for exclusion**

HW = handwashing

| **First author** | **Year** | **Title** | **Journal** | **Primary reason for exclusion** | **Comment** |
| --- | --- | --- | --- | --- | --- |
| Xiao | 1997 | Evaluation of effectiveness of comprehensive control for diarrhoea diseases in rural areas of east Fujian and analysis of its cost-benefit | Chinese Journal of Preventive Medicine | wrong intervention (HW <25% of content) | Broad intervention including water supply, sanitation, road renovation, ditch cleaning, health education (boil water, handwashing, fly prevention), advice to farmers (fences for poultry / livestock) |
| Uhari | 1999 | An open randomized controlled trial of infection prevention in child day-care centers. | Pediatric Infectious Disease Journal | not an economic evaluation | Does not report cost-effectiveness estimates, nor even costs |
| Guinan | 2002 | The effect of a comprehensive handwashing program on absenteeism in elementary schools | American Journal of Infection Control | partial economic evaluation | Measures cost savings only, does not value benefits |
| Mascie-Taylor | 2003 | The cost-effectiveness of health education in improving knowledge and awareness about intestinal parasites in rural Bangladesh | Economics and Human Biology | another reporting of an included study | Duplicates methods/results of Mascie-Taylor 1999 |
| Cimiotti | 2004 | A cost comparison of hand hygiene regimens | Nursing economics | wrong setting (healthcare, workplace, etc.) | Healthcare setting (but also reports costs not cost-effectiveness) |
| Hutton | 2007 | Global cost-benefit analysis of water supply and sanitation interventions | Journal of Water and Health | wrong intervention (0% or unclear HW) | listed interventions do not include hygiene/handwashing |
| Haller | 2007 | Estimating the costs and health benefits of water and sanitation improvements at global level | Journal of Water and Health | wrong intervention (0% or unclear HW) | listed interventions do not include hygiene/handwashing |
| Renwick | 2007 | Cost -Benefit Analysis of National and Regional Integrated Biogas and Sanitation Programs in Sub -Saharan Africa | Discussion paper | investment case for planned programme | Planning for a specific biogas, sanitation and hygiene programme |
| Edmond | 2010 | New Approaches to Preventing, Diagnosing, and Treating Neonatal Sepsis | PLoS Medicine | not an economic evaluation | Review article |
| Graves | 2012 | Evaluating the economics of the Australian National Hand Hygiene Initiative | Healthcare Infection | wrong setting (healthcare, workplace, etc.) | Healthcare setting (but also reports costs not cost-effectiveness) |
| Chen | 2013 | Cost-effectiveness of influenza control measures: A dynamic transmission model-based analysis | Epidemiology and Infection | partial economic evaluation | Estimates unit cost per person per year, not cost-effectiveness |
| Graves | 2013 | Linking scientific evidence and decision making: A case study of hand hygiene interventions | Infection Control and Hospital Epidemiology | wrong setting (healthcare, workplace, etc.) | Healthcare setting (but also reports costs not cost-effectiveness) |
| Zhang | 2013 | Promoting clean hands among children in Uganda: A school-based intervention using 'tippy-taps' | Public Health | partial economic evaluation | Reports costs only |
| Ataniya-zova | 2014 | A Cost-Benefit Analysis of Early Childhood Hygiene Interventions in Uzbekistan | Eurasian Journal of Business and Economics | partial economic evaluation | Measures cost savings only, does not value benefits as per our definition of benefit-cost analysis |
| Khazeni | 2014 | Health and Economic Benefits of Early Vaccination and Nonpharmaceutical Interventions for a Human Influenza A (H7N9) Pandemic | Annals of Internal Medicine | wrong intervention (HW <25% of content) | Intervention is closures of schools and child care facilities; home isolation; cough etiquette; hand washing; use of alcohol-based hand gels; and facemasks. Also unclear whether it is specifically promotion of handwashing or merely the practice of handwashing |
| Whinnery | 2016 | Handwashing with a water-efficient tap and low-cost foaming soap: the Povu Poa "Cool Foam" system in Kenya. | Global Health: Science and Practice | partial economic evaluation | Reports costs only |
| Cunning-ham | 2017 | Community video: An adaptable and effective tool for nutrition social and behavior change | Annals of Nutrition and Metabolism | not a study (conference abstract, opinion, etc.) | Conference abstract |
| OECD | 2018 | Stemming the Superbug Tide: Just A Few Dollars More | OECD report | wrong setting (healthcare, workplace, etc.) | Healthcare setting |
| Senapati | 2019 | A cholera metapopulation model interlinking migration with intervention strategies - a case study of zimbabwe (2008-2009) | Journal of Biological systems | another reporting of an included study | Duplicates methods/results of Sardar 2013 |
| Asamoah | 2020 | Global stability and cost-effectiveness analysis of COVID-19 considering the impact of the environment: using data from Ghana | Chaos Solitons & Fractals | wrong intervention (not promotion or provision) | Intervention is the practice of "cover coughs/ sneezes, wash hands after coughs/sneezes" rather than the promotion of that practice, per the description but also the costs. |
| Bagepally | 2021 | Cost-effectiveness of surgical mask, N-95 respirator, hand-hygiene and surgical mask with hand hygiene in the prevention of COVID-19: Cost effectiveness analysis from Indian context | Clinical Epidemiology and Global Health | wrong intervention (not promotion or provision) | Intervention is the practice of handwashing rather than the promotion of that practice, and costs only include "hand wash"/sanitiser |
| Zafari | 2021 | The cost-effectiveness of common strategies for the prevention of transmission of SARSCoV- 2 in universities | PLoS ONE | wrong intervention (not promotion or provision) | Intervention is the practice of handwashing rather than the promotion of that practice. In any case, handwashing and masking was the comparator ("status quo arm") rather than an intervention. |
| Asamoah | 2021 | Sensitivity assessment and optimal economic evaluation of a new COVID-19 compartmental epidemic model with control interventions | Chaos, Solitons and Fractals | wrong intervention (HW <25% of content) | Intervention is physical distancing, media advocacy, wearing of a nose mask, the use of hand sanitiser-washing of hands, lockdowns, stringent safety measures in hospitals (and/or isolation centres), with a constant supply of effective personal protective equipment (PPE)), testing-diagnoses and contact tracing. So it is also unclear if soap or sanitisier, and the intervention is practice rather than promotion. |
| Reddy | 2021 | Cost-effectiveness of public health strategies for COVID-19 epidemic control in South Africa: a microsimulation modelling study | Lancet Global Health | wrong intervention (not promotion or provision) | Handwashing is not one of the strategies tested |
| Wang | 2022 | Cost-Effectiveness of Public Health Measures to Control COVID-19 in China: A Microsimulation Modeling Study | Frontiers in Public Health | wrong intervention (not promotion or provision) | Intervention is the practice of handwashing rather than the promotion of that practice, per the description but also the included costs. |
| Akinyemi | 2023 | A tale of two countries: Optimal control and cost-effectiveness analysis of monkeypox disease in Germany and Nigeria | Healthcare Analytics | wrong intervention (not promotion or provision) | Intervention is "Behavioral modification and personal hygiene" and involves practice of handwashing rather than its promotion. |
| Edward | 2024 | On the role of vaccination, health education, and hygiene compliance in the elimination and control of Hepatitis A Virus: An optimal control approach | Informatics in Medicine Unlocked | wrong intervention (not promotion or provision) | Not clear what "hygiene" is and whether it includes handwashing, nor what is included in "health education". No cost information provided so unclear whether handwashing is part of it or not. |
| Omura | 2024 | Promoting healthy practices among schools and children in rural bangladesh: a randomised controlled trial of skill-based health education | BMC public health | partial economic evaluation | Cost study only because it is cost per 0.1 SD increase in the average treatment effect, where outcomes are behaviour (not health or other valued outcome) |
| Ssemanda | 2025 | Cost-effectiveness of interventions toward improving microbial food safety of chicken meat along supply chains in Burkina Faso and Ethiopia | International Journal of Food Microbiology | wrong intervention (not promotion or provision) | Cost itemisation in Supplementary Material makes it clear there is no promotion (Ethiopia study is in homes not workplace) |
| Engida | 2025 | Analysis of a Comprehensive Mathematical Model for Leptospirosis Dynamics: An Optimal Control Application | JOURNAL OF MATHEMATICS | wrong intervention (not promotion or provision) | Nothing about the control option suggests that handwashing is being promoted and no cost details provided to allow understanding of that. |

### F. Further details of included studies

**Table F‑1: Further characteristics of included studies**

| **Reference** | **Sensitivity analysis** | **Perspective** | **Comparator and behaviour** | **CHEERS**  **score (%)** |
| --- | --- | --- | --- | --- |
| Varley (1998) | one-way DSA on 1 parameter | Provider (but unclear who is bearing operational costs) | No intervention (unclear HW behaviour) | 66% |
| Mascie-Taylor (1999) | None, but reports results for all children and a subset with high-intensity worm infections | Provider implied (household-borne costs excluded) | No intervention (unclear HW behaviour) | 55% |
| Borghi (2002) | one-way DSA on 8 parameters | Societal (but provider and household perspectives also reported) | No intervention (“before” scenario behaviour) | 79% |
| Larsen (2003) | None | Societal implied (soap/water costs included) | Unclear – appears no intervention (unclear HW behaviour) | 40% |
| Cairncross (2006) | None | Provider implied (facility/soap/water costs excluded) | Unclear – appears no intervention (unclear HW behaviour) | 45% |
| Hansen (2008) | Unclear low/high scenarios for HWWS, plus one-way DSA on few variables for overall package of 65 interventions | Provider (household-borne costs excluded) | Unclear – appears current practice, which is not stated (unclear HW behaviour) | 59% |
| Lachance (2010) | One-way and two-way DSA on many variables, as well as scenario-based DSA | Societal | Thermometer provision only (control arm) but meta-analysis effect estimate used (self-reported HW behaviour) | 90% |
| Machdar (2013) | None | Unclear, appears provider | Unclear – appears no intervention (unclear HW behaviour) | 36% |
| Sardar (2013) | Unclear. Aspires to PSA given the nature of the model but methods are not explained. Distributions of key variables are not provided | Unclear, appears provider | Unclear – appears no intervention (unclear HW behaviour) | 54% |
| Siu (2021) | PSA with cost-effectiveness plane and cost-effectiveness acceptability curve. However, no DSA and not 95% CI for the ICER. | Societal | Promotion of water use in domestic vegetable gardening (control arm), with baseline/control behaviour reported | 82% |
| Azor-Martinez (2021) | Unclear. Claims Bayesian PSA but methods and distributions are not reported | Societal and provider | Usual handwashing practice (control arm), and sanitiser arm. Behaviour not reported. | 93% |
| Beresniak (2023) | DSA for pandemic scenarios reported for all interventions. Additionally PSA for certain interventions only (not HWWS) with unclear methods and distributions | Health system | Unclear – appears no intervention (unclear HW behaviour) | 55% |
| Whittington (2012) | PSA with uniform distributions for key variables, alongside DSA on adoption/adherence variables | Societal implied (default for BCA) | No intervention (uptake clear, but not baseline level of behaviour) | 81% |
| Larsen (2016) | Two-way DSA on discount rate and health valuation methods | Societal implied (default for BCA) | Unclear – appears no intervention (unclear HW behaviour) | 55% |
| Townsend (2017) | None for BCR. A 95% CI for net benefit is reported but methods for this are unclear. | Provider implied (only includes programme cost, not facility/soap/water/) but also includes avoided cost of illness on benefit side which is inconsistent. | Unclear – appears no intervention (assumptions about baseline behaviour reported) | 41% |

### G. CHEERS scores

Figure G‑0‑1: Distribution of CHEERS scores per item

% of studies (n=15)

|  | 1 (fully met) | 0.5 (partially met) | 0 (not met) | n/a |
| --- | --- | --- | --- | --- |
| 1. Title | 80% | 6.66667% | 13.3333% | 0% |
| 2. Abstract | 40% | 26.6667% | 13.3333% | 20% |
| 3. Objectives | 53.3333% | 40% | 6.66667% | 0% |
| 4. Population | 53.3333% | 40% | 6.66667% | 0% |
| 5. Setting | 13.3333% | 53.3333% | 33.3333% | 0% |
| 6. Perspective | 46.6667% | 26.6667% | 26.6667% | 0% |
| 7. Comparators | 20% | 60% | 20% | 0% |
| 8. Time horizon | 20% | 53.3333% | 26.6667% | 0% |
| 9.Discounting | 26.6667% | 33.3333% | 33.3333% | 6.66667% |
| 10. Outcomes | 93.3333% | 0% | 6.66667% | 0% |
| 11. Effectiveness | 40% | 33.3333% | 26.6667% | 0% |
| 12. Preference-based outcomes | 20% | 13.3333% | 13.3333% | 53.3333% |
| 13. Costs | 26.6667% | 46.6667% | 26.6667% | 0% |
| 14. Currencies & dates | 53.3333% | 13.3333% | 20% | 13.3333% |
| 15. Model choice | 33.3333% | 40% | 26.6667% | 0% |
| 16. Assumption transparency | 53.3333% | 26.6667% | 20% | 0% |
| 17. Method transparency | 26.6667% | 66.6667% | 6.66667% | 0% |
| 18. Parameter transparency | 33.3333% | 60% | 6.66667% | 0% |
| 19. Incremental costs & outcomes | 40% | 33.3333% | 26.6667% | 0% |
| 20. Uncertainty | 13.3333% | 26.6667% | 60% | 0% |
| 21. Sub-group heterogeneity | 20% | 20% | 0% | 60% |
| 22. Limitations | 33.3333% | 60% | 6.66667% | 0% |
| 23. Funding | 80% | 0% | 0% | 20% |
| 24. Conflicts of interest | 26.6667% | 0% | 0% | 73.3333% |

Note. Items are not applicable when norms are not established for that type of publication (e.g. abstract, funding, conflicts of interest) or the study did not use that method (e.g. preference-based outcomes, heterogeneity).

Figure G‑0‑2: CHEERS item scores and overall ratings and scores per study

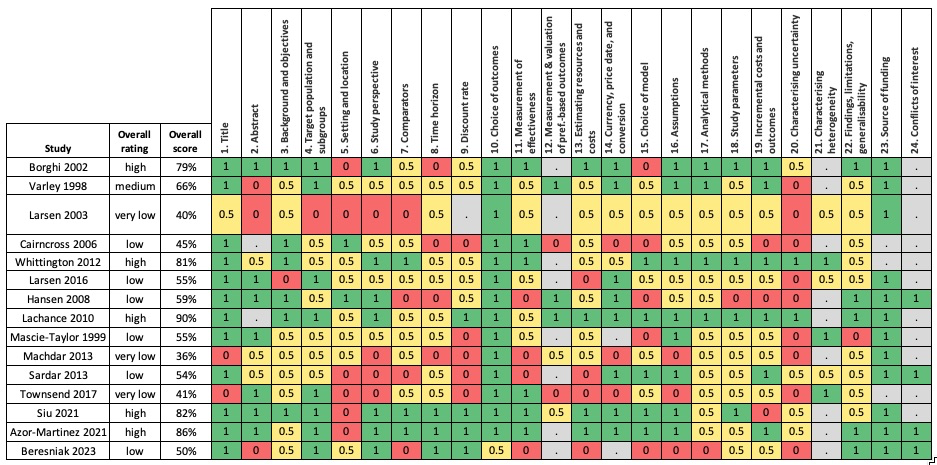

Figure G‑0‑3: CHEERS score by year of publication

CHEERS score (%)

| Year of publication | score |
| --- | --- |
| 1998 | 65.9091% |
| 1999 | 54.7619% |
| 2002 | 78.5714% |
| 2003 | 40.4762% |
| 2006 | 45% |
| 2008 | 58.6957% |
| 2010 | 90.4762% |
| 2012 | 80.9524% |
| 2013 | 36.3636% |
| 2013 | 54.3478% |
| 2016 | 54.7619% |
| 2016 | 40.9091% |
| 2021 | 81.8182% |
| 2021 | 86.3636% |
| 2023 | 50% |

### H. PRISMA 2020 checklist

| **Section and Topic** | **Item #** | **Checklist item** | **Location where item is reported** |
| --- | --- | --- | --- |
| **TITLE** | | |  |
| Title | 1 | Identify the report as a systematic review. | Title |
| **ABSTRACT** | | |  |
| Abstract | 2 | See the PRISMA 2020 for Abstracts checklist. | The abstract reports PRISMA 2020 for Abstracts items, to the extent possible within word limit. |
| **INTRODUCTION** | | |  |
| Rationale | 3 | Describe the rationale for the review in the context of existing knowledge. | Introduction |
| Objectives | 4 | Provide an explicit statement of the objective(s) or question(s) the review addresses. | Introduction |
| **METHODS** | | |  |
| Eligibility criteria | 5 | Specify the inclusion and exclusion criteria for the review and how studies were grouped for the syntheses. | Methods: Selection criteria |
| Information sources | 6 | Specify all databases, registers, websites, organisations, reference lists and other sources searched or consulted to identify studies. Specify the date when each source was last searched or consulted. | Methods: Search strategy |
| Search strategy | 7 | Present the full search strategies for all databases, registers and websites, including any filters and limits used. | Supplementary Material A |
| Selection process | 8 | Specify the methods used to decide whether a study met the inclusion criteria of the review, including how many reviewers screened each record and each report retrieved, whether they worked independently, and if applicable, details of automation tools used in the process. | Methods: Search strategy |
| Data collection process | 9 | Specify the methods used to collect data from reports, including how many reviewers collected data from each report, whether they worked independently, any processes for obtaining or confirming data from study investigators, and if applicable, details of automation tools used in the process. | Methods: Data extraction |
| Data items | 10a | List and define all outcomes for which data were sought. Specify whether all results that were compatible with each outcome domain in each study were sought (e.g. for all measures, time points, analyses), and if not, the methods used to decide which results to collect. | Methods: Selection criteria |
|  | 10b | List and define all other variables for which data were sought (e.g. participant and intervention characteristics, funding sources). Describe any assumptions made about any missing or unclear information. | Methods. Full extraction sheet is in the protocol (available via link in manuscript). |
| Study risk of bias assessment | 11 | Specify the methods used to assess risk of bias in the included studies, including details of the tool(s) used, how many reviewers assessed each study and whether they worked independently, and if applicable, details of automation tools used in the process. | Methods: Study quality |
| Effect measures | 12 | Specify for each outcome the effect measure(s) (e.g. risk ratio, mean difference) used in the synthesis or presentation of results. | Methods: Selection criteria |
| Synthesis methods | 13a | Describe the processes used to decide which studies were eligible for each synthesis (e.g. tabulating the study intervention characteristics and comparing against the planned groups for each synthesis (item #5)). | Methods: Data synthesis |
|  | 13b | Describe any methods required to prepare the data for presentation or synthesis, such as handling of missing summary statistics, or data conversions. | n/a (narrative) |
|  | 13c | Describe any methods used to tabulate or visually display results of individual studies and syntheses. | Methods: Data synthesis |
|  | 13d | Describe any methods used to synthesize results and provide a rationale for the choice(s). If meta-analysis was performed, describe the model(s), method(s) to identify the presence and extent of statistical heterogeneity, and software package(s) used. | Methods: Data synthesis |
|  | 13e | Describe any methods used to explore possible causes of heterogeneity among study results (e.g. subgroup analysis, meta-regression). | Methods: Data synthesis |
|  | 13f | Describe any sensitivity analyses conducted to assess robustness of the synthesized results. | Methods: Data synthesis (i.e. different thresholds) |
| Reporting bias assessment | 14 | Describe any methods used to assess risk of bias due to missing results in a synthesis (arising from reporting biases). | n/a – formal methods not possible for systematic reviews of economic evaluations |
| Certainty assessment | 15 | Describe any methods used to assess certainty (or confidence) in the body of evidence for an outcome. | n/a – GRADE and similar not applicable to systematic reviews of economic evaluations |
| **RESULTS** | | |  |
| Study selection | 16a | Describe the results of the search and selection process, from the number of records identified in the search to the number of studies included in the review, ideally using a flow diagram. | Figure 1 |
|  | 16b | Cite studies that might appear to meet the inclusion criteria, but which were excluded, and explain why they were excluded. | Supplementary Material E |
| Study characteristics | 17 | Cite each included study and present its characteristics. | Table 1 |
| Risk of bias in studies | 18 | Present assessments of risk of bias for each included study. | Table 2 |
| Results of individual studies | 19 | For all outcomes, present, for each study: (a) summary statistics for each group (where appropriate) and (b) an effect estimate and its precision (e.g. confidence/credible interval), ideally using structured tables or plots. | n/a – systematic review of economic evaluations, not effectiveness studies |
| Results of syntheses | 20a | For each synthesis, briefly summarise the characteristics and risk of bias among contributing studies. | Results |
|  | 20b | Present results of all statistical syntheses conducted. If meta-analysis was done, present for each the summary estimate and its precision (e.g. confidence/credible interval) and measures of statistical heterogeneity. If comparing groups, describe the direction of the effect. | n/a – no meta-analysis |
|  | 20c | Present results of all investigations of possible causes of heterogeneity among study results. | n/a – no meta-analysis |
|  | 20d | Present results of all sensitivity analyses conducted to assess the robustness of the synthesized results. | n/a – no meta-analysis |
| Reporting biases | 21 | Present assessments of risk of bias due to missing results (arising from reporting biases) for each synthesis assessed. | n/a – see above |
| Certainty of evidence | 22 | Present assessments of certainty (or confidence) in the body of evidence for each outcome assessed. | n/a – see above |
| **DISCUSSION** | | |  |
| Discussion | 23a | Provide a general interpretation of the results in the context of other evidence. | Discussion |
|  | 23b | Discuss any limitations of the evidence included in the review. | Discussion |
|  | 23c | Discuss any limitations of the review processes used. | Discussion |
|  | 23d | Discuss implications of the results for practice, policy, and future research. | Discussion |
| **OTHER INFORMATION** | | |  |
| Registration and protocol | 24a | Provide registration information for the review, including register name and registration number, or state that the review was not registered. | Abstract and methods |
|  | 24b | Indicate where the review protocol can be accessed, or state that a protocol was not prepared. | Methods and link in “Data sharing” sub-section |
|  | 24c | Describe and explain any amendments to information provided at registration or in the protocol. | n/a |
| Support | 25 | Describe sources of financial or non-financial support for the review, and the role of the funders or sponsors in the review. | Abstract and “Funding” sub-section |
| Competing interests | 26 | Declare any competing interests of review authors. | “Declaration of interests” sub-section |
| Availability of data, code and other materials | 27 | Report which of the following are publicly available and where they can be found: template data collection forms; data extracted from included studies; data used for all analyses; analytic code; any other materials used in the review. | Extraction sheet within linked protocol, data and code in “Data sharing” sub-section. No code used. |
